# Predicting parental Human papillomavirus vaccine hesitancy: development and internal validation of machine learning models

**DOI:** 10.64898/2026.09.18.26363393

**Authors:** Israel Oluwamayomikun Alabi, Daniel Biftu Bekalo, Christiana Kartsonaki, Valirie Ndip Agbor

## Abstract

**Objectives:** Cervical cancer is Cameroon’s second most common cancer among women. However, high parental Human papillomavirus (HPV) vaccine hesitancy impedes elimination, and no validated predictive tool exists. This study aimed to develop and internally validate machine learning models predicting parental HPV vaccine hesitancy in the Buea Health District and identify key predictors.

**Methods:** This secondary analysis used cross-sectional survey data from 1,156 parents of children aged 9–18 years. Hesitancy was derived from self-reported vaccination status and intention. Twenty-six predictor groups were screened via group LASSO (17 retained for logistic regression; all 26 for tree-based models). Data were split 80/20 into training (n=925) and test (n=231) sets. Logistic regression, random forest, and XGBoost were tuned via repeated cross-validation while thresholds were set by Youden’s J statistic. Discrimination, calibration, and Brier score were assessed on the test set. Predictions were interpreted using SHAP.

**Results:** All models showed comparable discrimination (AUC-ROC 0.841–0.870). XGBoost had the highest AUC (0.870, vs 0.869 for logistic regression, DeLong p=0.965); logistic regression had the highest sensitivity (73.1%), F1-score (74.9%), and lowest Brier score (0.1435). Calibration slopes exceeded 1 (1.12–1.46) across models. Insufficient vaccine information, perceived vaccine unsafety, and distrust in the Ministry of Health were the dominant predictors.

**Discussion:** The three models achieved comparable, clinically meaningful discrimination, and algorithmic complexity did not improve prediction over standard regression. Hesitancy was more strongly predicted by information access and institutional trust than sociodemographic disadvantage.

**Conclusion:** These findings support trust-building engagement over broad demographic campaigns.

**What is already known on this topic:** Parental HPV vaccine hesitancy is a major barrier to HPV vaccination, but predictive modelling evidence from sub-Saharan Africa is limited and no internally validated model for parental HPV vaccine hesitancy has been developed in the region.

**What this study adds:** We developed and internally validated logistic regression, random forest and XGBoost models for predicting parental HPV vaccine hesitancy, with all three showing comparable discrimination (AUC-ROC 0.841–0.870) and logistic regression achieving the lowest Brier score and highest sensitivity and F1-score. Insufficient vaccine information, perceived vaccine unsafety and distrust in the Ministry of Health were the dominant predictors across the models and interpretability analyses.

**How this study might affect research, practice or policy:** The findings support the development and external validation of simple, interpretable prediction tools to identify parents who may benefit from targeted, trust-building vaccine communication and community engagement to support HPV vaccination and cervical cancer prevention.

## Introduction

Cervical cancer remains the fourth most common cancer among women globally, with approximately 660,000 new cases and 350,000 deaths recorded in 2022 (1). Roughly 94% of these deaths occur in low- and middle-income countries (LMICs), where access to preventive services is severely limited (1,2). The World Health Organization (WHO) established the “90-70-90” targets, aiming for 90% of girls to be fully vaccinated against HPV by age 15 by 2030 (3,4). While vaccination confers approximately 95% protection against cervical cancer, achieving these targets remains a significant challenge in sub-Saharan Africa (SSA) (1,5,6).

In Cameroon, cervical cancer is the second most prevalent cancer among women, with an estimated 2,770 new cases and 1,787 deaths annually (7,8). The HPV vaccine was added to the national immunisation programme in October 2020; however, misinformation regarding infertility and adverse effects have reduced public acceptance and uptake (2,9). Parents play a critical role in improving HPV vaccine uptake among children because they largely determine whether their children are vaccinated. However, parental vaccine hesitancy remains substantial, with nearly half (46.8%) of parents in Buea reported to be vaccine hesitant (10). Evidence indicates that HPV vaccination and awareness are influenced by sociodemographic circumstances, with lower educational attainment associated with poorer HPV awareness and greater vaccine hesitancy among parents. Gender-related socioeconomic barriers may also limit access to preventive health services, highlighting the potential for existing inequalities to influence HPV vaccination decisions (9–11).

Previous studies in this context have identified population-level correlates of hesitancy using inferential logistic regression, without developing internally validated predictive tools for individual-level risk stratification (2,10). While logistic regression is effective for identifying broad associations, it often struggles to capture the complex, non-linear interactions and high-dimensional patterns inherent in diverse survey data without extensive manual feature engineering (12). Globally, machine learning (ML) methods are increasingly applied to complex, high-dimensional survey data to support public health decision-making, with algorithms such as random forest and XGBoost achieving up to 93% predictive accuracy for parental hesitancy in high-income settings (13). By automatically detecting intricate, multivariable interactions, ML algorithms can optimise predictive performance over standard regression models, making them better suited for generating precise individual-level risk scores (14,15). Comparative ML studies using locally collected HPV hesitancy data, however, remain limited in SSA, and most existing models are trained on data from western populations, limiting generalisability to non-similar settings (16,17).

This study developed and compared predictive models for parental HPV vaccine hesitancy, and by applying SHAP-based interpretability to translate model output into actionable public health insight. Such predictive insights are essential in developing culturally adapted communication strategies to assist the government and health workers in overcoming local resistance and support Cameroon’s effort towards the WHO’s global targets for cervical cancer elimination.

## Methods

### Study design and population

This was a secondary analysis of data from a community-based, cross-sectional survey conducted in the Buea Health District (BHD), South West Region, Cameroon. The health district is constituted of seven health areas: Bokwango, Bova, Buea Road, Buea Town, Molyko, Muea, and Tole (10). Data were collected between August 2023 and March 2024 using a modified version of the WHO standard tool for assessing vaccine hesitancy among adults (18).

Parents of children aged 9–18 years residing in the BHD were recruited via a two-stage sampling method: four health areas (Bokwango, Bueatown, Molyko, Muea) were randomly selected, followed by convenience sampling at community centres and households (10). Transient visitors and those who withdrew consent during the period of the study were excluded (10). The dataset comprised 1,187 participants.

### Outcome definition

The primary outcome, parental HPV vaccine hesitancy, was coded as a binary variable (1 = hesitant, 0 = acceptant), derived from a two-step response process. Parents were first asked whether their child had received the HPV vaccine (“Yes”/“No”/“Not sure”). Participants responding “No” or “Not sure” were then asked about their intention to vaccinate; a response of “No” or “Not sure” to this follow-up question was defined as vaccine hesitancy. This definition was applied uniformly across all participants and sociodemographic groups.

Outcome ascertainment was based on self-report and did not require subjective interpretation by an assessor; blinding of outcome assessment was therefore not applicable.

### Predictor variables

Predictors were organised into four domains: 1) sociodemographic (age, sex, health area, marital status, education, employment, household income); 2) knowledge and awareness (HPV and cervical cancer awareness, perceived disease severity, personal contact with a cervical cancer case); 3) attitudinal and behavioural (perceived vaccine safety, concern about adverse effects, trust in the Ministry of Health and pharmaceutical companies, vaccination history, chronic illness history); and 4) sociocultural (religious affiliation, perceived religious leader support, perceived peer vaccination behaviour).

No predictors required subjective interpretation by an assessor; blinding of predictor measurement to outcome/other predictors was therefore not applicable.

### Data preprocessing

Preprocessing was performed using R (v4.5.2). One duplicate record was removed, missingness was minimal (<5%) (Supplementary Table 1), and complete-case analysis yielded an analytic sample of *n*=1,156. The dataset was randomly partitioned into an 80% training set (n = 925) and a 20% held-out test set (n = 231) via stratified random sampling on the outcome, preserving the 53.2%/46.8% class distribution in both subsets. The held-out test set was reserved for final model evaluation and was not used for feature selection, model fitting, hyperparameter tuning, or threshold selection to avoid information leakage. An aliasing check performed on the training set using Variance Inflation Factor (VIF) (19) revealed perfect collinearity between general awareness of cervical cancer/HPV and specific knowledge regarding their severity and causes. Hence, the redundant variables were removed. One further variable (heard bad news about the vaccine) was dropped for near-zero variance. No significant outliers requiring transformation were identified. Sparse or conceptually overlapping categorical levels were collapsed (e.g., married/cohabiting; divorced/widowed; no formal/primary education; Islam/None religion merged into “Other”; “Not sure” responses grouped with “No” for select binary items) and categorical predictors were one-hot encoded for model matrix construction. The outcome distribution (53.2% acceptant, 46.8% hesitant) was sufficiently balanced that no resampling was needed.

### Feature selection

Predictors were grouped into 26 conceptual blocks (matching dummy levels of multi-level factors, or individual continuous variables) and selected using group LASSO regression, which selects entire factor groups jointly rather than isolated dummy levels, preserving interpretability (20). The regularisation parameter λ was selected using 10-fold cross-validation on the training data, with the value corresponding to the minimum cross-validated deviance. The logistic regression model was trained using the 17 predictors selected by group LASSO, whereas the random forest and XGBoost models were trained using all 26 predictors.

### Model development

Logistic regression, random forest, and XGBoost were implemented using standard model specifications (full equations in Supplementary Methods 1), and classification followed a decision threshold τ selected as described below.

#### Training and hyperparameter optimisation

With 541 hesitancy events and 17 retained predictor groups, the events-per-variable ratio (∼32) exceeds commonly cited minimum thresholds for stable logistic regression estimation, providing post hoc justification that the study size was adequate for model development (21). No prospective sample size calculation was performed, as this was a secondary analysis of an existing dataset. Each algorithm was tuned within the training set using repeated 10-fold cross-validation (3 repetitions). For random forest, a grid search over mtry ∈ {2, …, 26} maximised cross-validated area under the receiver-operating characteristic (AUC-ROC) curve. For XGBoost, hyperparameters were tuned using 5-fold cross-validation with early stopping based on AUC-ROC. The final model used a learning rate (η) of 0.05, maximum tree depth of 4, minimum child weight of 5, γ of 0.5, and row and column subsampling ratios of 0.8. For each model, the classification threshold was selected using predictions generated from the training data through cross-validation. Youden’s J statistic was used to identify the threshold that maximised the sum of sensitivity and specificity (22). The selected thresholds were then fixed before evaluating the models on the held-out test set. The selected thresholds were 0.461 for logistic regression, 0.513 for random forest, and 0.476 for XGBoost.

### Model evaluation

Model performance was assessed on the independent test set using accuracy, sensitivity, specificity, and F1-score, with vaccine-hesitant parents defined as the positive class. Discrimination was assessed using the area under the receiver operating characteristic curve (AUC-ROC) with 95% confidence intervals. Overall probabilistic prediction accuracy was assessed using the Brier score. Calibration was assessed using the calibration intercept and slope with 95% confidence intervals and visually using calibration plots (reliability diagrams) comparing mean predicted probabilities with observed event proportions across deciles of predicted risk in the held-out test set. Definitions and formulas for all performance metrics are provided in Supplementary Methods 2.

Fairness and subgroup performance were assessed by evaluating discrimination and calibration across prespecified sociodemographic and geographic subgroups, including sex, education level, and health area, to identify potential disparities or heterogeneity in model performance across population groups.

### Model interpretation

Feature importance was extracted from tree-based models using Mean Decrease in Impurity (random forest) and Gain (XGBoost) (full definitions in Supplementary Methods 3). Although they identify global feature rankings, they cannot capture the direction of individual predictor effects.

To capture both magnitude and direction of predictor effects, SHAP (SHapley Additive exPlanations) values were computed from the final XGBoost model on the held-out test set (n=231) to reflect generalisable feature contributions (formula in Supplementary Methods 3).

## Results

### Sample characteristics

Of the 1,156 participants included in the final analysis, 46.8% (*n* = 541) were classified as vaccine-hesitant and 53.2% (*n* = 615) as acceptant. Baseline characteristics stratified by hesitancy status are presented in Table 1 (Full details in Supplementary Table 2).

**Table 1:**
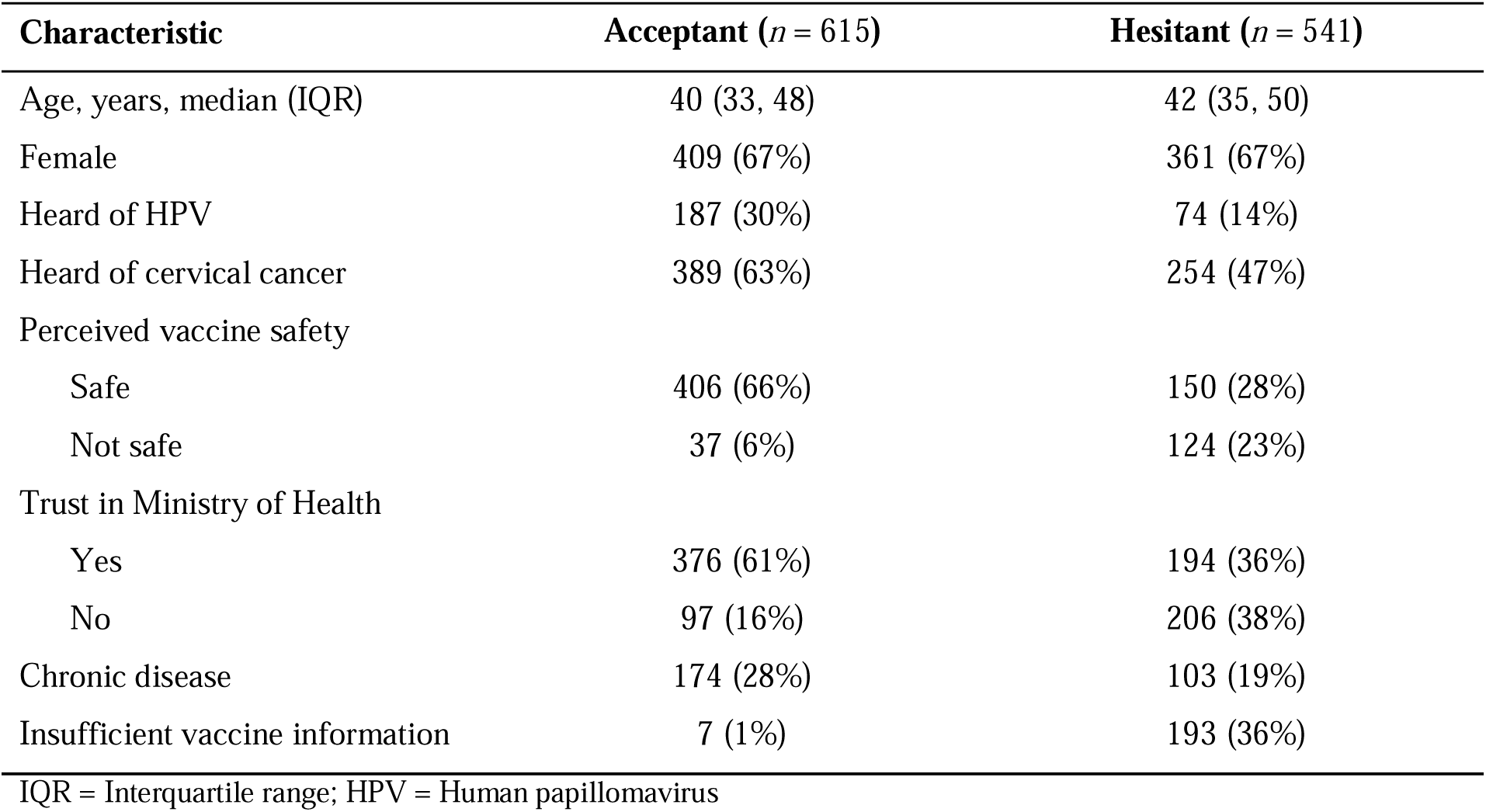
Selected characteristics of study participants by vaccine hesitancy status.

### Hyperparameter optimisation

The logistic regression model, fitted on 17 group-LASSO-selected predictor groups, had a residual deviance of 865.74 on 898 degrees of freedom (AIC = 919.74), compared with a null deviance of 1278.56 on 924 degrees of freedom. Grid search identified mtry = 6 as optimal for the random forest, using 500 trees and repeated 10-fold cross-validation (3 repetitions), with AUC-ROC as the tuning criterion. For XGBoost, cross-validated early stopping identified η = 0.05, maximum depth = 4, γ = 0.5, minimum child weight = 5, and row/column subsampling of 0.8 (Supplementary Table 3).

### Model performance

All three models achieved high accuracy (0.766–0.775) and comparable discrimination (AUC-ROC 0.841-0.870) on the held-out test set (n=231; prevalence=46.8%; Table 2). Logistic regression achieved the highest sensitivity (73.1%) and F1-score (74.9%); random forest achieved the highest accuracy (77.5%) and specificity (82.9%); XGBoost achieved the highest AUC (0.870) (Figure 1A). XGBoost significantly outperformed random forest (DeLong p=0.016), but neither ensemble model significantly outperformed logistic regression (p>=0.053), indicating no detectable discrimination advantage from added nonlinear flexibility. Calibration slopes exceeded 1 for all models (1.12– 1.46), with near-zero intercepts, indicating mild underconfidence at the extremes of risk (Table 2). Logistic regression had the lowest Brier score (0.1435; Brier Skill Score 42.3% over the base rate). Followed by XGBoost (0.1482; 40.5%) and random forest (0.1627; 34.6%). Discrimination and calibration were broadly consistent across sex, education level and health area, with no evidence of meaningful subgroup disparities (Supplementary Table 4).

**Figure 1:**
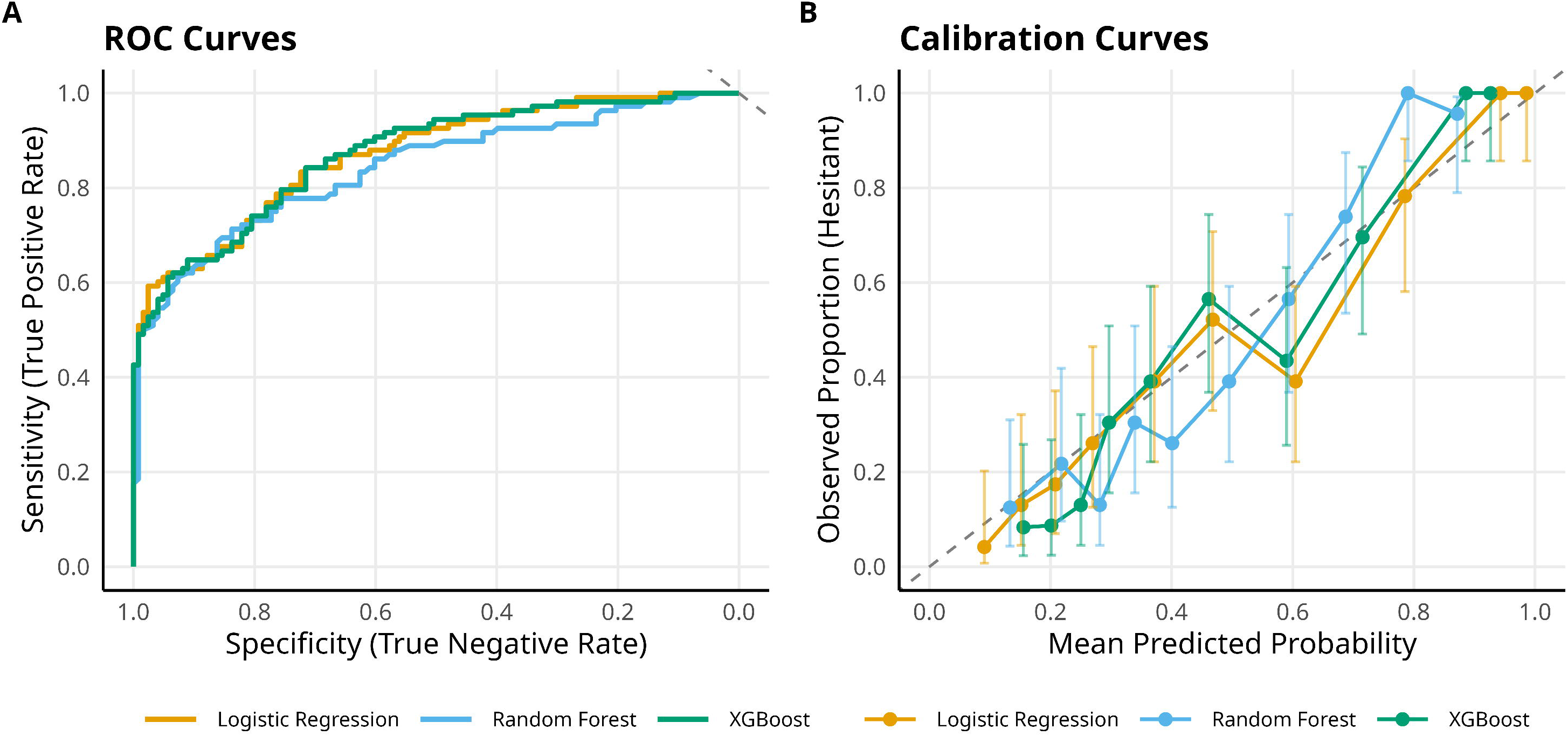
Discrimination and Calibration plots for models predicting parental HPV vaccine hesitancy

**Table 2:** Comparative performance of the prediction models on the held-out test set

| Performance metric | Logistic Regression | Random Forest | XGBoost |
| --- | --- | --- | --- |
| AUC-ROC | 0.869 (0.824–0.914) | 0.841 (0.789–0.893) | 0.870 (0.825–0.915) |
| Accuracy | 0.771 (0.711–0.823) | 0.775 (0.716–0.827) | 0.766 (0.706–0.819) |
| Sensitivity | 0.731 (0.638–0.812) | 0.713 (0.618–0.796) | 0.722 (0.628–0.804) |
| Specificity | 0.805 (0.724–0.871) | 0.829 (0.751–0.891) | 0.805 (0.724–0.871) |
| F1-score | 0.749 (0.680–0.809) | 0.748 (0.674–0.812) | 0.743 (0.673–0.807) |
| Brier score | 0.1435 | 0.1627 | 0.1482 |
| Calibration intercept | □0.124 (□0.458,<br>0.211) | □0.062 (□0.356,<br>0.232) | □0.089 (□0.397,<br>0.218) |
| Calibration slope | 1.124 (0.820–1.427) | 1.460 (1.084–1.837) | 1.438 (1.071–1.805) |

Values are presented as point estimates with 95% confidence intervals. AUC-ROC = area under the receiver operating characteristic curve.

### Predictor importance and SHAP explainability

Predictor importance was consistent across all three interpretability approaches. In the logistic regression model, insufficient vaccine information was the strongest predictor (OR = 35.05, 95% CI: 16.82–85.73, p<0.001), followed by perceived vaccine unsafety (OR = 4.85) or uncertainty (OR = 2.15), distrust in the Ministry of Health (OR = 2.17), never having heard of HPV (OR = 1.80), age (OR = 1.02/year), and chronic disease history (OR = 0.48) (Table 3).

**Table 3:**
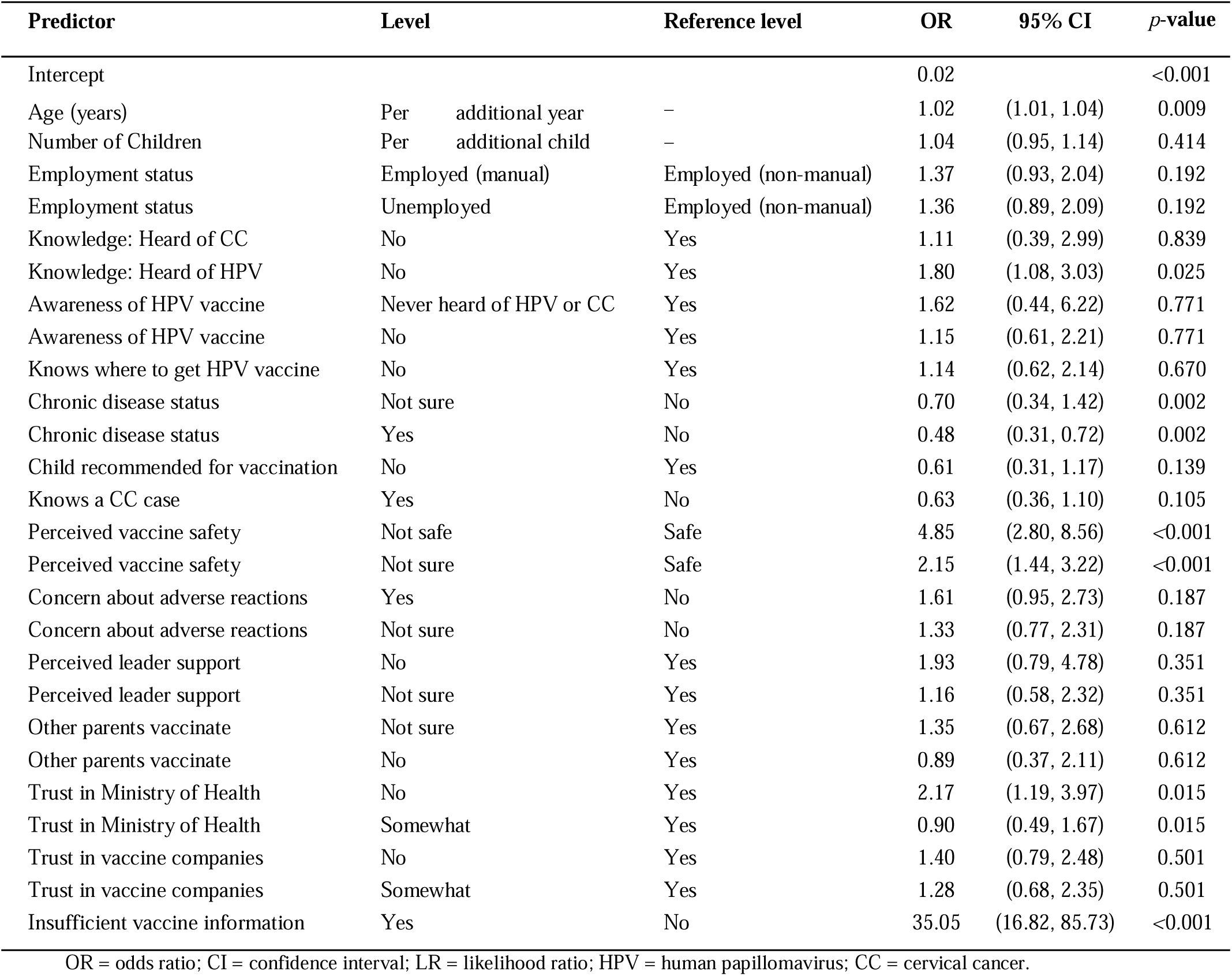
logistic regression model specification.

No complete or quasi-complete separation was detected by the formal diagnostic, although insufficient vaccine information showed an extreme prevalence imbalance that warranted diagnostic attention.

Random forest, XGBoost, and SHAP rankings converged on the same top predictors which are insufficient information, perceived safety, and institutional trust (Figure 2).

**Figure 2:**
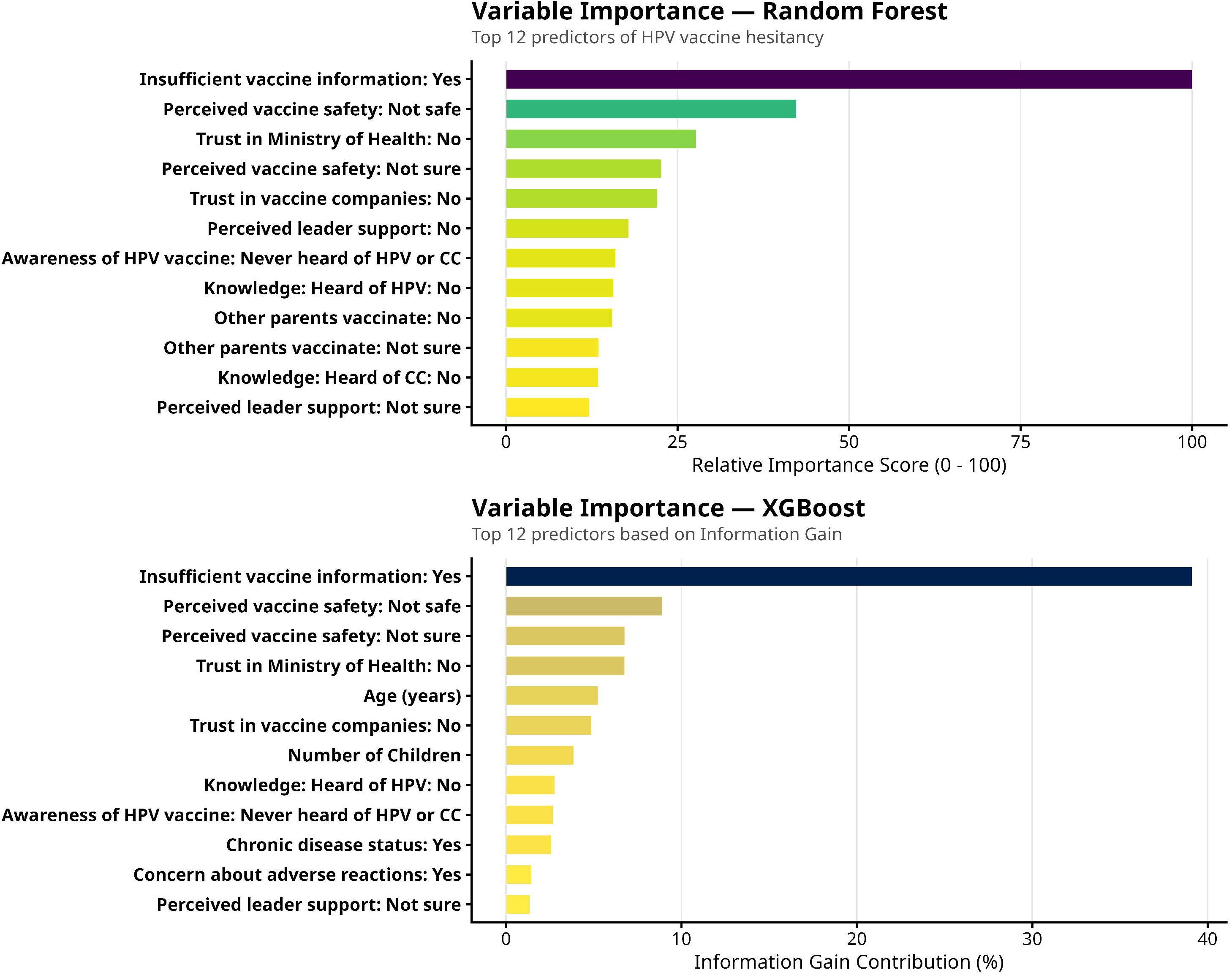
Top 12 predictors of HPV vaccine hesitancy in the random forest and XGBoost model

In addition, SHAP beeswarm plot (Supplementary Figure 1) confirms the direction of each effect. In particular, insufficient vaccine information, perceiving the vaccine as unsafe, uncertainty about vaccine safety, and lack of trust in the Ministry of Health or vaccine companies were associated with higher predicted hesitancy. Conversely, having a history of chronic disease had a protective SHAP contribution. SHAP waterfall plots for the least and most predicted cases (Supplementary Figure 2a and 2b) illustrated how individual-level factors contributed to predicted hesitancy.

SHAP dependence analysis (Supplementary Figure 3) revealed an “institutional trust paradox”: among parents reporting insufficient information, the hesitancy-driving effect was stronger among those who still trusted the Ministry of Health than among those who already distrusted it, and conversely, among distrustful parents, having sufficient information was associated with higher hesitancy risk than feeling uninformed. For safety perceptions, among parents who considered the vaccine unsafe, those who had sufficient information showed higher hesitancy risk than those without, a pattern that may be consistent with exposure to misinformation presented as official communication. Age exhibited a non-linear, sigmoidal relationship with hesitancy risk, protective from ages 20–40, with an inflection between ages 40–45 and a plateau of elevated risk (SHAP ≈ +02 to +0.4) after age 50, an effect associated with institutional distrust in older parents.

### Sensitivity analysis

A pre-specified sensitivity analysis using an alternative 70/30 train–test split (n=810/346) confirmed the stability of model discrimination (AUC-ROC 0.834–0.861; maximum absolute change from the primary analysis = 0.007) and predictor rankings, with insufficient vaccine information remaining the strongest predictor (OR = 32.00, 95% CI: 15.30–78.45; full results in Supplementary Table 5). One additional predictor, personally knowing someone with cervical cancer, was protective in this split (OR = 0.46, 95% CI: 0.24–0.85) but not in the primary analysis, likely reflecting sampling variability rather than a substantive difference.

## Discussion

This study aimed to develop and evaluate predictive models for parental HPV vaccine hesitancy in the BHD. To our knowledge, this is the first study to apply and compare multiple ML algorithms, incorporating group LASSO feature selection and SHAP-based interpretation, to a locally collected, community-based dataset on parental HPV vaccine hesitancy in sub-Saharan Africa.

Three principal findings emerge. First, predictive classification of hesitancy is feasible using survey-based data in this low-resource setting, with all three models achieving comparable, clinically meaningful discrimination. Second, logistic regression matched or exceeded the ensemble models on sensitivity, F1-score, and Brier score, and its AUC did not differ significantly from XGBoost’s, challenging the assumption that algorithmic complexity necessarily improves prediction in structured, moderate-sized health survey data. Third, predictor-importance rankings across all three models consistently favoured information access and institutional trust over socio-demographic variables such as educational attainment and income, suggesting these attitudinal/institutional factors may be more actionable intervention targets in this setting. The broadly consistent performance across sociodemographic and health-area subgroups is reassuring from a fairness perspective.

These findings align with Cameroonian and Nigerian studies identifying institutional distrust, low HPV awareness, and absent provider recommendation as dominant hesitancy drivers (2,23). Discrimination was somewhat lower than global ML benchmarks using larger, US-based datasets (AUC up to 0.93), (13) reflecting this study’s smaller, culturally distinct sample. Our findings are also consistent with reports that ensemble methods yield only marginal gains over logistic regression in structured health-survey data (24).

The findings in this study have implications for population health. The identified high-risk phenotype (older parents lacking HPV awareness, distrustful of the Ministry of Health, and reporting insufficient vaccine information) could inform a brief, SHAP-informed screening tool potentially deployable at routine well-child visits or community health meetings, enabling precision targeting in place of blanket communication campaigns, which have shown limited effectiveness in Cameroon (9,10). The dominance of information insufficiency as a predictor suggests that expanding the reach of information via community health workers, peer educators, and local radio may yield greater reductions in hesitancy than refining content for already-informed parents. Given the centrality of institutional distrust, community-based dialogic engagement leveraging traditional and religious leaders and drawing on the demonstrated success of door-to-door and Periodic Intensification of Routine Immunisation (PIRI) campaigns in the Southwest Region (9), is likely to be more effective than information provision alone. Finally, the protective association observed for chronic disease history suggests that existing points of healthcare contact represent an underused opportunity for integrated HPV vaccine counselling.

For potential implementation, predictor inputs should be checked against development-time coding. If a predictor is missing or invalid, the prediction should be flagged as unavailable rather than imputed ad hoc, and any future imputation strategy should first be validated. The tool is intended to support, not replace, clinical or public-health judgement. In a future implementation, data entry could be performed by trained healthcare or community health workers with basic digital literacy and without specialist ML expertise to inform, rather than dictate, targeted counselling.

Methodologically, this study demonstrates that group LASSO provided a parsimonious, group-preserving predictor set for the logistic regression model, while SHAP converted black-box XGBoost predictions into epidemiologically actionable contribution scores. The finding that logistic regression matched ensemble methods reinforces prior evidence cautioning against the uncritical adoption of complex ML models for structured, moderate-sized clinical and public health datasets (25).

Limitations include the cross-sectional design, which precludes causal inference; reliance on self-reported survey measures, which may be subject to social desirability bias; and the single-district sampling frame, which limits external generalisability to other regions of Cameroon.

## Conclusion

Logistic regression, random forest, and XGBoost achieved comparable and clinically meaningful discriminative ability, with logistic regression yielding the highest sensitivity and lowest Brier score. Across all models and interpretability methods, hesitancy was consistently predicted by insufficient vaccine information, perceived vaccine unsafety, and distrust in health authorities, indicating that hesitancy in this context is fundamentally a problem of information access and institutional trust rather than demographic disadvantage.

These findings support a shift from generic, top-down vaccination campaigns toward precision public health strategies that equip local community leaders and frontline healthcare workers to deliver targeted, trust-building engagement. Future work should externally validate these models in other regions of Cameroon, incorporate longitudinal data to clarify causal pathways, and investigate integration of model-derived risk scores into routine immunisation systems.

## Data Availability

All data produced in the present study are available upon reasonable request to the authors

## Declarations

### Ethics approval and consent to participate

Ethical approval for the primary data collection study was obtained from the Ethics Committee and Institutional Review Board of the University of Buea (Ethics Registration number 2023/1949-02). As this study is a secondary analysis of anonymised survey data, no additional ethical clearance was required. All participants in the primary study provided informed consent.

### Author contributions

IOA: Conceptualisation, data curation, formal analysis, visualization, writing – original draft. DBB: supervision and writing – review & editing. CK: Conceptualisation, supervision and writing – review & editing. VNA: Conceptualisation, supervision, writing – review & editing. All authors provided substantial input and approved the final version of the manuscript.

### Availability of data

De-identified individual participant data will be made available to researchers who provide a methodologically sound proposal, subject to approval from the University of Buea IRB, following publication. Requests should be directed to.

### Code availability

Analysis code is not currently publicly available; interested parties may contact the corresponding author.

### Protocol and registration

No study protocol was pre-registered, and this study was not registered in a clinical trials or study registry.

### Patient and public involvement

Patients and the public were not involved in the design, conduct, reporting, or dissemination of this study.

### Funding

There is no financial support for this study.

### Consent for publication

Not applicable

### Conflicts of interest

The authors declare that they have no conflicts of interest.

## Notes

### Competing Interest Statement

The authors have declared no competing interest.

### Author Declarations

The study was approved by the Ethics Committee and Institutional Review Board of the University of Buea (Ethics Registration number 2023/1949-02).

## References

1. World Health Organization. Cervical cancer [Internet]. [cited 2026 Aug 9]. Available from: https://www.who.int/news-room/fact-sheets/detail/cervical-cancer

2. Kehbila J, Mbinta JF, Ekabe CJ, Joselyne MN. Prevalence and Determinants of HPV Vaccine Hesitancy Among Parents in Efoulan Health District Yaoundé [Internet]. In Review; 2025 [cited 2026 Aug 9]. Available from: https://www.researchsquare.com/article/rs-7642911/v1 doi:10.21203/rs.3.rs-7642911/v1

3. World Health Organization. Global strategy to accelerate the elimination of cervical cancer as a public health problem [Internet]. [cited 2026 Aug 9]. Available from: https://www.who.int/publications/i/item/9789240014107

4. WHO African Region. WHO AFRO Investment Case Series: Accelerating Cervical Cancer Elimination in Africa through Strengthened HPV Vaccination, Screening and Treatment [Internet]. [cited 2026 Aug 9]. Available from: https://www.afro.who.int/publications/who-afro-investment-case-series-accelerating-cervical-cancer-elimination-africa

5. LaMontagne DS, Bloem PJN, Brotherton JML, Gallagher KE, Badiane O, Ndiaye C. Progress in HPV vaccination in low and lower middle income countries. Int J Gynecol Obstet. 2017 Jul;138(S1):7–14. doi:10.1002/ijgo.12186

6. Borda H, Bloem P, Akaba H, Guillaume D, Willens V, Jurgensmeyer M, et al. Status of HPV disease and vaccination programmes in LMICs: Introduction to special issue. Vaccine. 2024 Jul;42:S1–8. doi:10.1016/j.vaccine.2023.10.062

7. ICO/IARC HPV Information Centre. Cameroon: Human Papillomavirus and Related Cancers, Fact Sheet 2023. Fact Sheet. 2023.

8. Cheuyem FZL, Tchamani R, Bodo EML, Achangwa C, Dabou S, Adama M, et al. HPV prevalence and associated factors in Cameroon: a systematic review and meta-analysis [Internet]. Epidemiology; 2026 [cited 2026 Aug 9]. Available from: http://medrxiv.org/lookup/doi/10.64898/2026.02.15.26346335 doi:10.64898/2026.02.15.26346335

9. Njoh AA, Waheed DEN, Kedakse TSNJ, Ebongue LJ, Kongnyuy EJ, Amani A, et al. Overcoming challenges and achieving high HPV vaccination uptake in Cameroon: lessons learned from a gender-neutral and single-dose program and community engagement. BMC Public Health. 2025 May 8;25(1):1696. doi:10.1186/s12889-025-22776-3

10. Ntonifor MMN, Tazinkeng NN, Kemah BL, Claudia NE, Sonia YK, Nchinjoh SC, et al. Factors associated with parental hesitancy towards the human papillomavirus vaccine: a cross-sectional study. Sci Rep. 2025 May 26;15(1):18284. doi:10.1038/s41598-025-94067-1

11. Fru CN, Andrew T, Greenspan D, Cho FN, Martin M, Livingstone J, et al. Vaccination Hesitancy: The Case of Cervical Cancer Vaccination in Fako Division, Cameroon. Int J Trop Dis Health. 2021 Jun 15;32–43. doi:10.9734/ijtdh/2021/v42i830476

12. Shahid M, Yahya MA, Song J, Naveed HM, Yuksel S, Dincer H, et al. Machine learning vs. traditional logistic regression: predictive performance and risk factor identification for child nutritional outcome in Pakistan. BMC Public Health. 2025 Nov 25;26(1):667. doi:10.1186/s12889-025-25621-9

13. Zheng Y, Frew PM, Wang D, Song Y, Patterson-Lomba O, Feizi A, et al. Developing and validating machine learning models to predict vaccine hesitancy and literacy among adults in the United States. Front Public Health. 2026 Mar 11;14:1669058. doi:10.3389/fpubh.2026.1669058

14. Hu Y, Zhang X, Slavin V, Belsti Y, Tiruneh SA, Callander E, et al. Beyond Comparing Machine Learning and Logistic Regression in Clinical Prediction Modelling: Shifting from Model Debate to Data Quality. J Med Internet Res. 2025 Nov 5;27:e77721. doi:10.2196/77721

15. Maxey NJ, Griffin TS, Powla PP, Pabon-Rodriguez FM. Comparative evaluation of machine learning models for predicting COVID-19 vaccine uptake in U.S. adults. BMC Public Health. 2026 Apr 9;26(1):1625. doi:10.1186/s12889-026-27199-2

16. Betsch C, Schmid P, Heinemeier D, Korn L, Holtmann C, Böhm R. Beyond confidence: Development of a measure assessing the 5C psychological antecedents of vaccination. Angelillo IF, editor. PLOS ONE. 2018 Dec 7;13(12):e0208601. doi:10.1371/journal.pone.0208601

17. Chait RM, Nastiti A, Chintana DA, Sari PN, Marasabessy N, Firdaus MI, et al. Using the Social–Ecological Model to Assess Vaccine Hesitancy and Refusal in a Highly Religious Lower–Middle-Income Country. Int J Environ Res Public Health. 2024 Oct 9;21(10):1335. doi:10.3390/ijerph21101335

18. World Health Organization. Data for action: achieving high uptake of COVID-19 vaccines [Internet]. [cited 2026 Sep 4]. Available from: https://www.who.int/publications/i/item/WHO-2019-nCoV-vaccination-demand-planning-2021.1

19. Marquardt DW. Generalized Inverses, Ridge Regression, Biased Linear Estimation, and Nonlinear Estimation. Technometrics. 1970 Aug;12(3):591–612. doi:10.1080/00401706.1970.10488699

20. Meier L, Van De Geer S, Bühlmann P. The Group Lasso for Logistic Regression. J R Stat Soc Ser B Stat Methodol. 2008 Feb 1;70(1):53–71. doi:10.1111/j.1467-9868.2007.00627.x

21. Riley RD, Ensor J, Snell KIE, Harrell FE, Martin GP, Reitsma JB, et al. Calculating the sample size required for developing a clinical prediction model. BMJ. 2020 Mar 18;m441. doi:10.1136/bmj.m441

22. Youden WJ. Index for rating diagnostic tests. Cancer. 1950;3(1):32–5. doi:10.1002/1097-0142(1950)3:1<32::AID-CNCR2820030106>3.0.CO;2-3

23. Yusuf KK, Olorunsaiye CZ, Gadanya MA, Ouedraogo S, Abdullahi AA, Salihu HM. HPV vaccine hesitancy among parents and caregivers of adolescents in Northern Nigeria. Vaccine X. 2024 Dec;21:100591. doi:10.1016/j.jvacx.2024.100591

24. Zhao Y. Vaccine Hesitancy Prediction Based on Machine Learning Algorithms. Highlights Sci Eng Technol. 2024 Mar 13;85:1016–24. doi:10.54097/tw73y836

25. Christodoulou E, Ma J, Collins GS, Steyerberg EW, Verbakel JY, Van Calster B. A systematic review shows no performance benefit of machine learning over logistic regression for clinical prediction models. J Clin Epidemiol. 2019 Jun;110:12–22. doi:10.1016/j.jclinepi.2019.02.004

